# COVID-19 epidemic severity is associated with timing of non-pharmaceutical interventions

**DOI:** 10.1101/2020.09.15.20194258

**Authors:** Manon Ragonnet-Cronin, Olivia Boyd, Lily Geidelberg, David Jorgensen, Fabricia F. Nascimento, Igor Siveroni, Robert Johnson, Marc Baguelin, Zulma M Cucunubá, Elita Jauneikaite, Swapnil Mishra, Hayley A Thompson, Oliver Watson, Neil Ferguson, Christl A. Donnelly, Erik Volz, on behalf of the Imperial COVID-19 Response Team

**Affiliations:** MRC Centre for Global Infectious Disease Analysis and the Department of Infectious Disease Epidemiology, Imperial College London; Department of Statistics, University of Oxford

## Abstract

**Background:** Unprecedented public health interventions including travel restrictions and national lockdowns have been implemented to stem the COVID-19 epidemic, but the effectiveness of non-pharmaceutical interventions is still debated. International comparisons are hampered by highly variable conditions under which epidemics spread and differences in the timing and scale of interventions. Cumulative COVID-19 morbidity and mortality are functions of both the rate of epidemic growth and the duration of uninhibited growth before interventions were implemented. Incomplete and sporadic testing during the early COVID-19 epidemic makes it difficult to identify how long SARS-CoV-2 was circulating in different places. SARS-CoV-2 genetic sequences can be analyzed to provide an estimate of both the time of epidemic origin and the rate of early epidemic growth in different settings.

**Methods:** We carried out a phylogenetic analysis of more than 29,000 publicly available whole genome SARS-CoV-2 sequences from 57 locations to estimate the time that the epidemic originated in different places. These estimates were cross-referenced with dates of the most stringent interventions in each location as well as the number of cumulative COVID-19 deaths following maximum intervention. Phylodynamic methods were used to estimate the rate of early epidemic growth and proxy estimates of epidemic size.

**Findings:** The time elapsed between epidemic origin and maximum intervention is strongly associated with different measures of epidemic severity and explains 46% of variance in numbers infected at time of maximum intervention. The reproduction number is independently associated with epidemic severity. In multivariable regression models, epidemic severity was not associated with census population size. The time elapsed between detection of initial COVID-19 cases to interventions was not associated with epidemic severity, indicating that many locations experienced long periods of cryptic transmission.

**Interpretation:** Locations where strong non-pharmaceutical interventions were implemented earlier experienced much less severe COVID-19 morbidity and mortality during the period of study.

**Funding:** MRC is jointly funded by the UK Medical Research Council (MRC) and the UK Department for International Development (DFID) under the MRC/DFID Concordat agreement and this award is part of the EDCTP2 programme supported by the European Union (MR/R015600/1). RJ and EV acknowledge funding from the European Commission (CoroNAb 101003653). EV additionally acknowledges funding from the Wellcome Trust (220885/Z/20/Z).

## Introduction

To prevent the spread of the SARS-CoV-2 virus, countries have implemented unprecedented measures ranging from school closures and travel bans to large-scale lockdowns^1^. Because of the immense economic and social consequences of such interventions, the utility of travel restrictions and lockdowns have been questioned by the public and media. Mathematical models can help determine whether these “non-pharmaceutical interventions” (NPI) have been effective in reducing transmission. Previous investigations based on mathematical models have shown that travel restrictions in Wuhan delayed arrival of the virus into other Chinese cities by around 3 days, and early implementation of control measures was associated with lower incidence^2^. And in Germany, changes in epidemic growth rates are correlated with the timing of interventions^3^. In order to assess the impacts of NPIs across European countries, Flaxman et al. estimated the rate of epidemic growth over time from reported COVID-19 deaths and pooled information across European countries^4^. The impact of NPIs was significant across all countries, reducing the reproduction number below 1, and implied a substantial reduction in the number of deaths compared to a scenario with no intervention. Differences in case reporting complicate international comparisons, but as the scale of epidemics has varied so drastically across countries, such comparisons are essential. Dye et al.^5^ found that the 100-fold difference in cumulative COVID-19 deaths across European countries was not attributable to lower rates of transmission and epidemic growth, but rather due to differences in dates of national lockdown.

Comparisons beyond Europe are more complex due to greater variability in epidemiological surveillance and epidemic growth rates. Furthermore, while sustained transmission took place across most of Europe from around mid-February^6^, global comparisons are complicated by the fact that the virus was introduced at very different times in different places. Epidemiological models are typically fitted using data on reported infections, deaths, and seroprevalence surveys. These provide estimates of the basic reproduction number, R_0_, the average number of onward transmissions per case in a susceptible population; and the time-varying reproduction number, R_t_. However, because a high proportion of SARS-CoV-2 infections are not detected and seroprevalence surveys have only recently been carried out, parameterizing models for epidemics in February through April when large scale NPIs were enforced is not easy. Critically, it is usually not understood how long SARS-CoV-2 circulated in populations prior to detection which is essential information for comparing timing of NPIs between locations.

Viral genetic data provide an alternative source of information for understanding when epidemics originated and how quickly they grew. Analysis of SARS-CoV-2 genetic sequences can help distinguish between imported and local transmission; and inclusion of sample dates allows for further time-resolution of epidemic dynamics, yielding dates of viral introduction into specific regions. Phylodynamic analysis uses genetic data to parametrise epidemiological models, for example estimating R_t_ directly from viral sequences^7^. Analyses of viral sequences have demonstrated that transmission of SARS-CoV-2 went undetected within the USA from mid-January^8–11^ and many epidemics were seeded between states rather than as a result of international travel. In contrast in Guangdong, China, the majority of new diagnoses appeared to be imports, demonstrating the effectiveness of surveillance and interventions in interrupting community transmission^12^.

In the present analysis, we reconstruct the epidemic trajectories of SARS-CoV-2 outbreaks for locations across the world for which genetic data were available. We estimate the time of viral introduction into each region and calculate time to lockdown for each region. We then determine whether reported and estimated epidemic sizes relate to the duration of times to lockdown.

## Methods

### Genetic data preparation and selection of study sites

SARS-CoV-2 sequences were downloaded from GISAID (http://gisaid.org)^13^ on June 7th 2020. Genetic data were cleaned and prepared for time-resolved phylogenetic analysis in R v3.6.1 (Supplementary Methods). We chose to focus our analysis on sites with at least 100 sequences available (although some sites with fewer sequences were analysed, see Supplementary Methods for details). When data were available for sites located within each other (e.g. New Orleans in Louisiana), the smaller geographic unit was preferentially selected (see Supplementary Methods and Discussion). Sites known to comprise many sequences from travellers or generated as a result of contact tracing were excluded. Because our model assumes that samples are taken at random from the population, we excluded duplicate sequences from our analyses as a proxy for membership with the same transmission chain. We investigated the effect of this choice through simulation (see Supplementary Methods).

### Transmission model and comparative phylodynamic analysis

We utilized a compartmental structured coalescent model in the BEAST2 v6.1 PhyDyn package^14,15^ to estimate the effective reproduction number and the number of infections through time from SARS-CoV-2 genetic sequences. The compartmental model has been previously described^16–18^ and applied to SARS-CoV-2 sequence data. Importantly the model allows for bidirectional migration from and into an international reservoir, and splits the infected compartment into categories representing individuals with high or low rates of onward transmission. Earlier modeling efforts in SARS-CoV-1 and SARS-CoV-2 demonstrated that the inclusion of a high transmission rate compartment is crucial to realistically capturing case numbers^18,19^. The ability to accurately reconstruct epidemic dynamics using the Bayesian Markov chain Monte Carlo (MCMC) inferential framework and phylodynamic model was assessed in a simulation experiment (Supplementary Methods). For each site under investigation, we simultaneously reconstructed a phylogeny and estimated epidemiological parameters in BEAST2 (see Supplementary Methods for details). In order to expand our analysis to sites that did not converge in the Bayesian MCMC using default parameters, we additionally calculated viral effective population size through time for each site using a non-parametric skygrowth model^20,21^ (Supplementary Methods). Independently, we estimated the timing of viral introductions through time-resolved phylogenetic analysis and parsimony reconstruction (Supplementary Methods). From the distribution of viral introduction times, we calculated the mean time of viral introduction weighted by the number of samples descended from each viral introduction. We refer to this weighted mean as the central epidemic seed time (CEST).

### Comparison with other sources of data and statistical analysis

For sites analysed using BEAST, we examined the relationship between mobility data provided by Google (google.com/covid19/mobility, analysis limited to “transit stations” only) and R_t_ ^22^ by calculating Pearson’s correlation coefficient. Google mobility data are measured in relative deviations from maximum mobility prior to the WHO pandemic declaration. For every site, we extracted dates of lockdown from the Oxford COVID-19 Government Response Tracker^1^. If the region never underwent a full lockdown, we used the date of the most severe national intervention (Sup Tab 2), henceforth named maximum NPI. We obtained case and death counts for comparison with our own results and to assess case reporting (Sup Tab 2). We then calculated the time from epidemic origin (CEST) to maximum NPI and the time from the tenth reported case to maximum NPI for each site and constructed a series of linear regression models. Time delays were counted in days. For all sites, we built linear regression models with time from CEST to maximum NPI as predictor variable and number of reported cases at maximum NPI and number of deaths one month after the maximum NPI as outcomes, respectively. To evaluate if the time that cases were first reported could serve as a proxy for time of epidemic origin, we reran these models changing our predictor variable to time between the tenth reported case and maximum NPI. For BEAST locations, we built a univariate regression model with time from CEST to maximum NPI as a predictor variable and the estimated number of infections as an outcome. We then expanded all three CEST-based models into multivariable models, including population size and R_0_ as predictors. When results appeared sensitive to the presence of outliers, we verified associations using median-based linear models^23^ implemented in the *mblm* R package v0.12.1^24^. We computed Pearson correlation coefficients between reported and estimated infections among BEAST locations. Population size, effective viral population size, estimated infections, reported infections and death counts were log-transformed prior to analysis. All analyses were conducted in R v3.6.1.

## Results

We identified 30 locations (15 in Europe, six in North America, five in the Middle East, three in Asia and one in Africa) where dates of public health interventions could be obtained and where publicly available SARS-CoV-2 sequences enabled inference of epidemic origin dates and reproduction numbers using model-based phylodynamic methods. We further identified a larger set of 57 locations (24 in Europe, 20 in North America, five in the Middle East, six in Asia, one in South America and one in Africa) meeting inclusion criteria that could be analyzed using non-parametric phylodynamic methods. In total, 29,163 SARS-CoV-2 sequences were analysed, in regional datasets ranging from 23 sequences (Innsbruck, Austria) to 677 sequences (Denmark). Sample dates ranged from 2020-01-08 to 2020-05-30.

Phylogenetic analysis indicates that epidemic seeding was a continuous process with all regions showing evidence for more than one epidemic origin (Figure 1). The earliest seeding events among the 57 sites took place in January and in most locations seeding continued with a maximum frequency occurring in March before maximum NPI and travel restrictions were implemented in most locations. Sites with larger census population sizes had older central epidemic seeding times (CEST, see Methods; r^2^=0.20, p<0.001).

**Figure 1.**
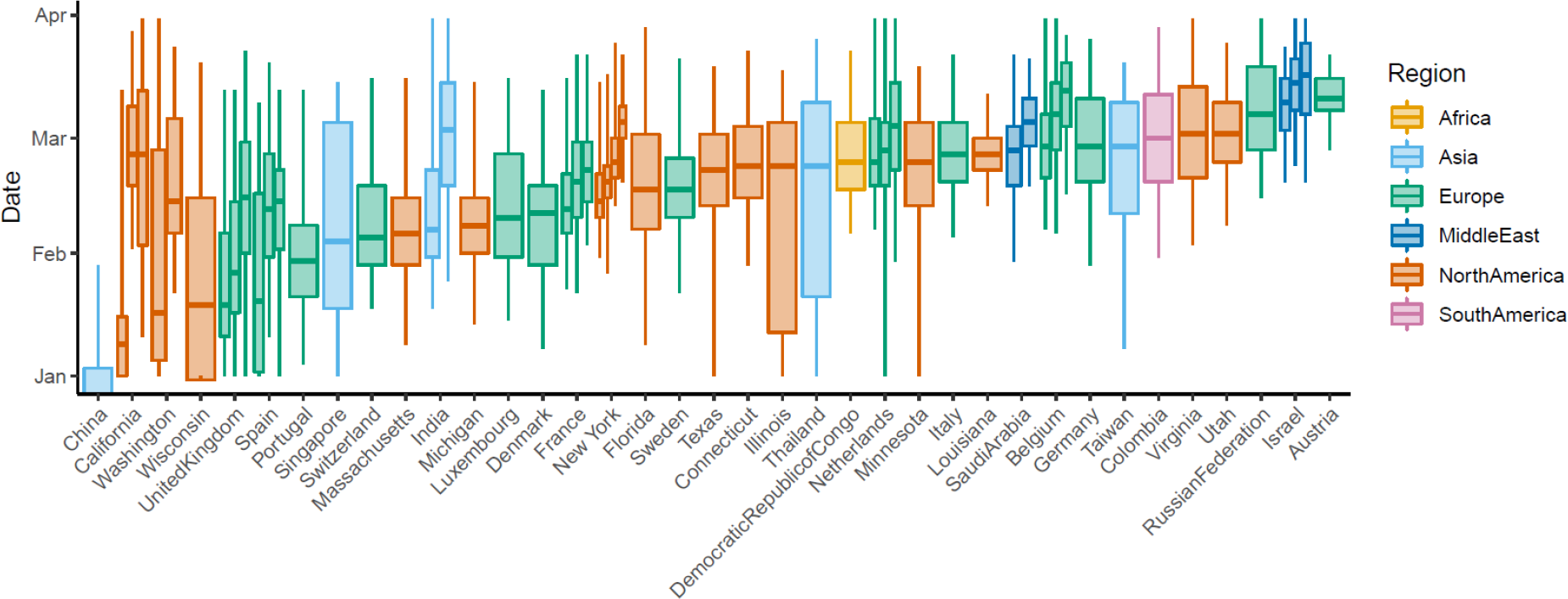
Distribution of phylogenetically-inferred seeding times for the 57 sites included in our analysis. Central box-plot lines represent the central epidemic seeding time (CEST). Boxes represent the interquartile range of the distribution. Sites from the same country or U.S state are grouped together and sites are ordered based on the first CEST for that country or U.S. state.

Among the 57 sites, CEST to maximum NPI was significantly associated with the number of reported infections at maximum NPI (p=3.8×10^−5^; Figure 2, Sup Fig 1, Sup Tab 1). This time delay explains over a quarter of the variance in the reported number of infections at time of maximum NPI (R^2^=0.27), and an additional 14 days of transmission before maximum NPI was associated with a 1.06 (95%CI: 0.59-1.54) log increase in number of reported infections. The time delay from the tenth reported case to maximum NPI was also predictive of the number of reported infections but explained less of its variance (R^2^=0.24, p=1.0×10^−4^). Census population size was a significant predictor of reported infections in univariate models (R^2^=0.17, p=1.3×10^−3^), but when included alongside time from CEST in multivariable models, it was no longer significant.

**Figure 2.**
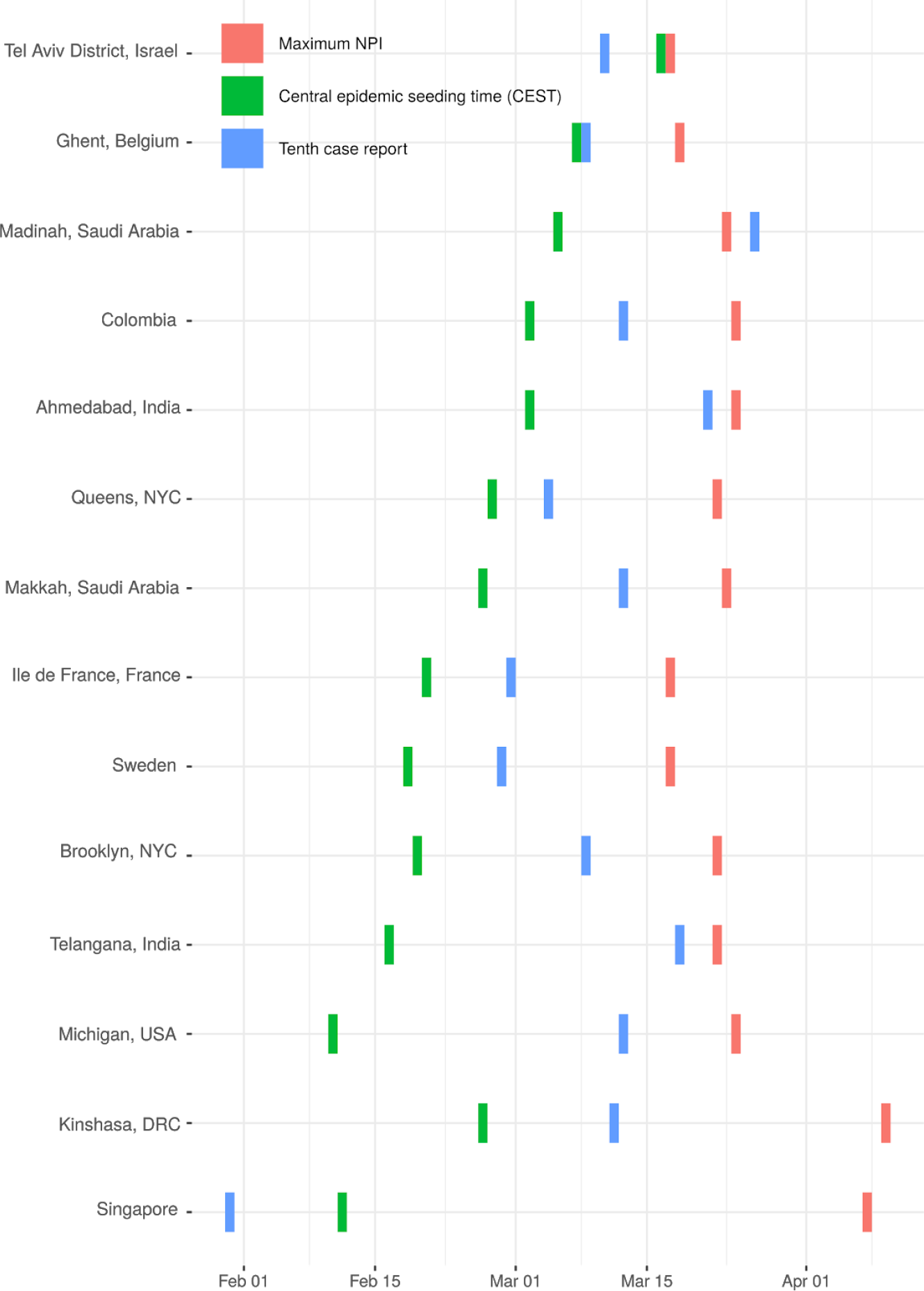
Dates of central epidemic seeding time (CEST), tenth case report and maximum non-pharmaceutical interventions (NPI) for 14 of 57 sites included in our analysis. Sites are ordered by the duration of time between CEST and maximum NPI. Note that the CEST does not represent the earliest viral introduction but rather the mean time of viral introduction weighted by the number of samples descended from each viral introduction; thus it can be preceded by the tenth diagnosis if early diagnoses are not phylogenetically related to later infections (e.g. Singapore). We selected up to three sites from each world region for which sites were available (Europe, North America, Middle East, Asia, Africa) based on their having the highest death counts in the region. Sup Fig 1 shows these dates for all 57 sites.

Among the 30 sites analysed using the Bayesian phylodynamic model, we generated estimates of the effective reproduction number R_0_, R_t_ and the number of infections over time (Sup Fig 3 and 4, Sup Tab 1 and 2). The time to maximum NPI was associated with the model-based estimates of the number of infections at maximum NPI (R^2^=0.19, p=1.7×10^−2^; Figure 3). An additional 14 days of transmission before maximum NPI was associated with a 0.60 (95%CI:0.11–1.08) log increase in the number of estimated infections at time of maximum NPI. In the multivariable model (R^2^ =0.44), time to maximum NPI remained significant (p=2.4×10^−3^). The estimated reproduction number R_0_ was also significant (p=7.0×10^−3^) as R_0_ is co-inferred alongside the number of infections in the model. A 25% increase in R_0_ was associated with a 0.90 (95%CI: 0.27-1.54) log increase in the number of estimated infections at maximum NPI.

**Figure 3.**
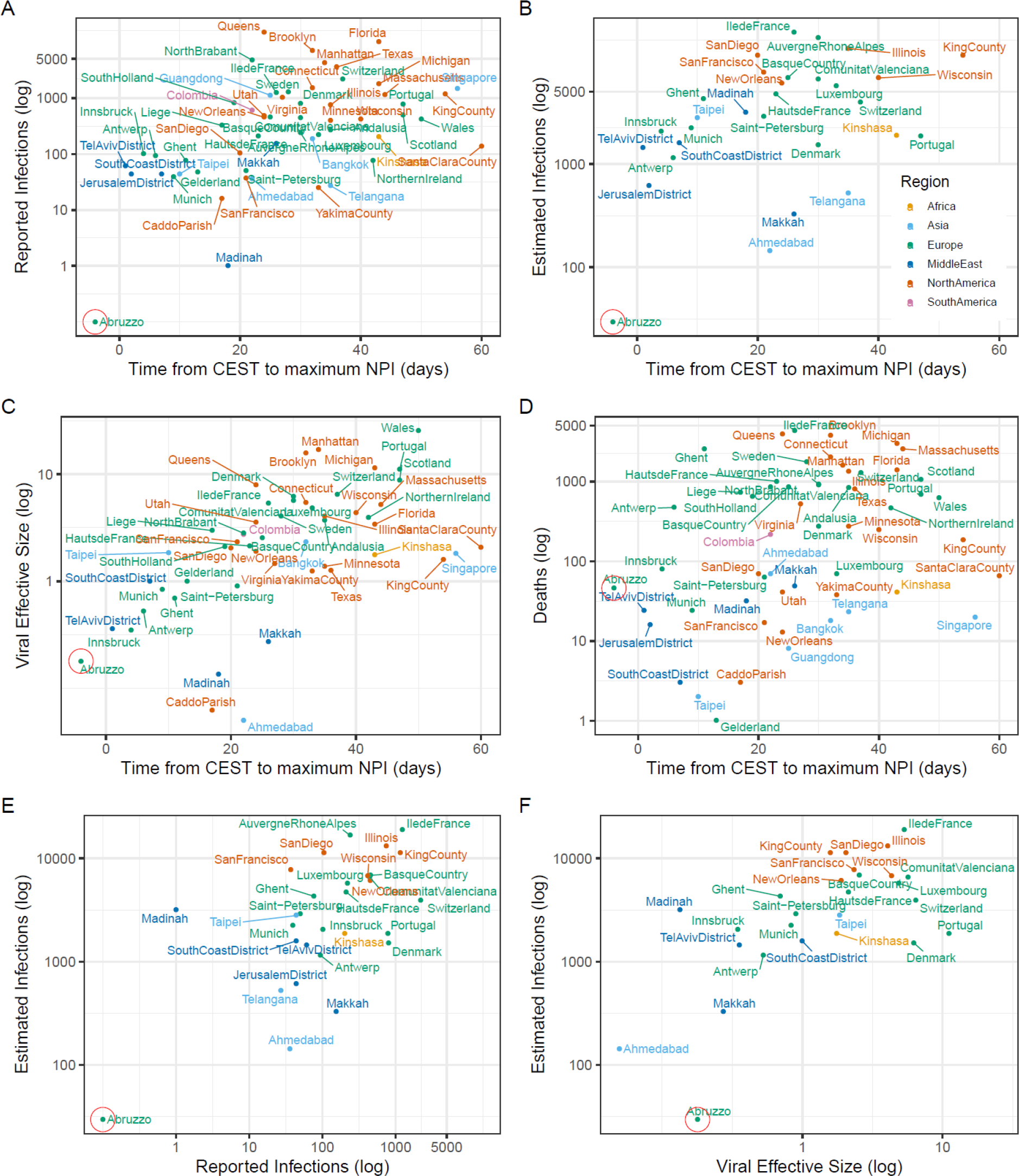
Relationship between different measures of epidemic severity and the estimated time to maximum NPI. A) Reported diagnosed infections versus time to max NPI. B) Phylodynamic estimates of cumulative infections at time of max NPI. C) Viral effective population size versus time to max NPI. D) Cumulative deaths within a month following max NPI. E) Phylodynamic estimate of infections versus reported diagnosed cases at time of max NPI. F) Estimated infections at max NPI versus virus effective population size at max NPI. Abruzzo is circled because it was an outlier possibly driving our correlations. As such, we repeated all analyses excluding Abruzzo.

We observed the same associations using non-parametric inference methods. Viral effective population size at maximum NPI, a proxy for the number of infections, was associated with time to maximum NPI (R^2^=0.32, p=1.2×10^−5^; Figure 3, Sup Tab 1). An additional 14 days of transmission before maximum NPI was associated with a 0.73 (95%CI:0.43-1.04) log increase in effective population size at time of maximum NPI. In a univariate model, census population size was predictive of viral effective size (R^2^=0.15, p=4.6×10^−3^). In a multivariable model (R^2^=0.38), time from CEST to maximum NPI remained a significant predictor (p=5.4×10^−4^) but census population size did not (p>0.05).

As an alternative measure of epidemic severity, we also considered the number of deaths reported at each site one month following the time of maximum NPI (Sup Fig 4, Sup Tab 1 and 2). The number of deaths was correlated with the estimated number of infections at maximum NPI (30 BEAST sites only, Pearson’s r=0.39, p=3.2×10^−2^; Figure 3)and with viral effective population size (all 57 sites, Pearson’s r=0.58, p=5.2×10^−6^; Figure 3). Time from CEST to maximum NPI was predictive of the number of deaths one month later (R^2^=0.11, p=1.1×10^−2^), but time from the tenth case was not (p>0.05). An additional 14 days of transmission before maximum NPI was associated with a 0.69 (95%CI:0.17-1.20) log increase in deaths one month after maximum NPI. Census population size of each location was not significant in a model predicting deaths (p>0.05).

Across all sites and using non-parametric methods, we estimated the mean epidemic doubling time was 3.68 days (IQR: 2.45–5.76). Across 30 sites analysed in BEAST using model-based phylodynamic methods, doubling time was 3.47 days (IQR: 3.09-4.83). Estimates of R_0_ varied between 1.53 (in Makkah, Saudi Arabia) and 4.18 (in New Orleans, Louisiana, USA; Sup Tab 3; Sup Fig 3). We did not detect an association between R_0_ and climatological variables consisting of mean rainfall and temperature over the period of uninhibited growth (results not shown).

To corroborate inference of effective reproduction numbers and epidemic size from genetic data, we examined the relationship between inferred quantities and reported numbers of cases as well as independent information about human mobility patterns. We found that changes in the inferred reproduction number through time, R_t_, corresponded with changes in Google human mobility metrics for 29 of 30 locations where these data were available (Sup Tab 3; Sup Fig 2). Large reductions in human mobility metrics consistently correspond with periods when R_t_ decreases. The mean cross-correlation between R_t_ and mobility metrics among those 29 locations was 0.79, ranging from 0.48 to 0.93 (all p<0.001). We then examined the relationship between the number of inferred cumulative infections at maximum NPI and the number of reported infections at that point. These were highly correlated (Pearson’s r=0.64, p=1.5×10^−4^; Figure 3). Using phylodynamic estimates of the cumulative number infected we estimated that the mean reporting rate (proportion of infections diagnosed) at the time of maximum NPI was 11.1% but varied greatly between regions and over time (Sup Tab 3).

We examined the sensitivity of the results to choice of phylodynamic methodology and compared inferred numbers infected to non-parametric estimates of viral effective population size. Estimates of size were highly correlated (Pearson’s r=0.67, p=1.4×10^−4^), but estimates of R_0_ were not (p>0.05). Finally, we conducted a sensitivity analysis by excluding our data point for “Abruzzo, Italy” (circled in Figure 3) in case it was driving observed associations. All relationships were maintained except the regression of estimated number of infections over time from CEST to intervention, which was no longer significant (Figure 3, panel B). However, when we applied a median based linear model^23^ to the dataset, the association between the estimated number of infections and time from CEST to intervention remained significant whether or not Abruzzo was excluded (p=8.9×10^−4^and p=7.1×10^−5^, respectively).

## Discussion

Among 57 geographical sites sampled across five continents, we found that time from SARS-CoV-2 introduction to time of lockdown (or maximum non pharmaceutical intervention [NPI] in locations that never underwent a full lockdown) was associated at a significance level of 0.001 with the severity of the epidemic in each location. This result is consistent with analyses^25^ conducted thus far separately in China^2,26^ and Europe^3–5^ and demonstrates the importance of cryptic transmission before testing and interventions were implemented in exacerbating epidemic severity. Notably, the time between detection of the tenth case at each site at the maximum NPI was not predictive of the number of deaths. This indicates that many locations experienced long and variable periods of cryptic transmission before epidemics were detected ^8–11^. This analysis implies that although outcomes are highly variable, on average implementing a strong NPI such as national lockdown two weeks earlier would approximately halve cumulative deaths in the period immediately following lockdown.

A previous comparison across European countries found that earlier lockdown dates were associated with fewer deaths, and that countries with fewer COVID-19 deaths had fewer inhabitants^5^. An association between population size and the number of deaths suggests that the depletion of susceptibles limited onward spread in places with smaller populations. However, serological surveys and patterns of per-capita death do not support the hypothesis that herd immunity has been reached anywhere in Europe^27^. In our global analysis, census population size was predictive of the number of estimated infections, viral effective population size and the number of deaths in univariate models. But when controlling for the time from CEST to lockdown, population size no longer had an effect, which is consistent with the hypothesis that herd immunity has not substantially limited transmission.

International comparisons of NPI effectiveness have been complicated by widely varying testing strategies in different locales and most epidemiological models are highly reliant on reported COVID-19 diagnoses and deaths. Our ability to run analyses on such a wide range of locations derives from the fact that our model is parameterised entirely by genetic data. We have shown that this approach produces accurate estimates of the numbers of infections in simulations (Supplementary Methods), and among the study sites, we found that the number of estimated infections was highly correlated with the numbers of COVID-19 diagnoses and deaths. Our analyses further support the use of viral effective population size at maximum NPI as a proxy for SARS-CoV-2 epidemic size. Where possible, we compared our estimates to those modelled or measured elsewhere. Estimates of R_0_ and R_t_ were in line with those previously reported for the same locations^28–32^ and R_t_ showed a strong association with changes in human mobility over time, as previously demonstrated for R_t_ estimates derived from traditional epidemiological models^22^. Dates of introduction into Europe aligned with previous reports demonstrating sustained transmission from mid-February^6^.

A limitation of our analysis is that the Bayesian MCMC for our phylodynamic model did not converge for all the locations. This can occur because one of the model assumptions is violated; for example, samples may not be collected at random, or the population is not randomly mixing. We addressed these concerns by excluding sites known to have prioritised sequencing from travellers^16,33^ or contact tracing and by focusing on smaller geographical units, such as cities and small regions where within-sample geographic structure is less biasing. Further optimisation of some parameters, such as the parameter for transmission overdispersion could have improved estimates, as has been recently demonstrated by Miller et al.^16^, but we chose to keep this parameter constant across sites to facilitate meta-analysis.

The non-parametric phylodynamic analysis allowed us to include sites for which data were available but the Bayesian MCMC did not converge^20,21^ as well as to examine sensitivity of results to choice of modelling framework. Results echoed those from the compartmental phylodynamic model. Viral effective population size at maximum NPI was associated with deaths one month after maximum NPI and with time to maximum NPI.

In conclusion, we have shown that across five continents, longer delays from viral introduction to lockdowns have led to more infections at lockdown and more deaths one month after lockdown. Our models were calibrated entirely using genetic data and thus provide an independent confirmation of mathematical models calibrated to traditional data sources. These findings emphasize the importance of NPI for decreasing epidemic severity, reinforce previous findings that seroprevalence is far below that needed for herd immunity ^27^ and highlight the risk for re-emergence and continued transmission.

## Data Availability

All genetic data are publicly available upon registration with GISAID (https://www.gisaid.org/). Details for non-genetic data are provided in our Supplementary Materials.

## Supplementary Figure Legends

Supplementary Figure 1: Dates of central epidemic seeding time (CEST), tenth case report and maximum non-pharmaceutical interventions (NPI) for all 57 sites included in our analysis. Sites are ordered by the duration of time between CEST and maximum NPI.

Supplementary Figure 2: Correlation between the effective reproduction number R_t_ (in purple) and changes in mobility over time (obtained from Google and measured in relative deviations from maximum mobility prior to the WHO pandemic declaration. The Pearson’s correlation coefficient is calculated only on the time period up until the last sample for that site. Mobility data were not available for Kinshasa, DRC.

Supplementary Figure 3: Cumulative estimated infections, estimated daily infections and effective reproduction number through time (R_t_) for all 30 locations analyzed in BEAST. Reported infections for each location are shown using yellow dots, while estimated infections are represented by black lines with 95% highest posterior density estimates.

Supplementary Figure 4: Reported number of infections at maximum NPI, reported number of deaths one month after maximum NPI for 57 sites included in the analysis. Estimated number of infections at maximum NPI are displayed for 30 sites analysed using BEAST. Sources for reported numbers are available in Sup Tab 2.

Supplementary Figure 5: Cumulative estimated infections, estimated daily infections and effective reproduction number through time (R_t_) for five simulated datasets. Analysis results are shown each time for the dataset analysed as whole and in deduplicated form. True values are shown using yellow dots and reconstructed estimates are shown as black lines with 95% highest posterior density estimates.

